# Antibody response to SARS-CoV2 among COVID-19 confirmed cases, and correlates with neutralizing assay in a subgroup of patients in Delhi National Capital Region, India

**DOI:** 10.1101/2022.05.17.22275193

**Authors:** Puneet Misra, Shashi Kant, Randeep Guleria, Sanjay Rai, Abhishek Jaiswal, Suprakash Mandal, Guruprasad R Medigeshi, Mohammad Ahmad, Anisur Rahman, Meenu Sangral, Kapil Yadav, Mohan Bairwa, Partha Haldar, Parveen Kumar

## Abstract

**Background:** Plaque reduction neutralization test (PRNT) is the gold standard to detect neutralizing capacity of the serum antibodies. Neutralizing antibody confers protection against further infection. The present study was done with the objective to measure the antibody level against SARS-CoV2 among laboratory confirmed COVID-19 cases and to evaluate whether the presence of anti-SARS-CoV2 antibodies indicate virus neutralizing capacity.

**Methods:** One hundred COVID-19 confirmed cases were recruited. Sociodemographic details and history of COVID-19 vaccination, contact with positive COVID-19 cases, and symptoms were ascertained using a self-developed semi-structured interview schedule. Serum samples of the participants were collected within three months from date of the positive report of COVID-19. The presence of anti-SARS-CoV-2 antibodies (IgA, IgG and IgM antibodies), receptor binding domain antibodies (anti-RBD), and neutralizing antibodies were measured.

**Findings:** Almost all participants had Anti-SARS-CoV2 antibodies (IgA, IgG and IgM) (99%) and Anti-RBD IgG antibodies (97%). However, only 69% had neutralizing antibodies against SARS-CoV2. Anti-RBD antibody levels were significantly higher among participants having neutralizing antibodies compared to those who didn’t.

**Interpretation:** The present study highlights that presence of antibodies against SARS-CoV2, or presence of anti-RBD antibody doesn’t necessarily imply presence of neutralizing antibodies.

**Funding:** World Health Organisation

## Introduction

Corona Virus disease (COVID-19) is caused by the virus SARS-CoV-2, which is a single-stranded RNA virus belonging to the genus Betacoronavirus. (1) It emerged in Wuhan, China in December 2019, and was declared as a global pandemic by WHO on 11 March 2020.

The SARS-CoV-2 infection causes a wide range of clinical manifestations ranging from cough, fever and malaise to severe pneumonia and acute respiratory distress syndrome. (2,3) Antibody mediated (humoral immunity) immunity is thought to play a vital role in the protection both in naturally infected and vaccinated people. The SARS-CoV-2 virus induces a classic antibody response in which IgM antibodies appear first followed by IgG antibodies which remain detectable for several months post-symptom onset (PSO) while IgM declines by 2-3 weeks of PSO. (4) Various serological tests are available to detect these antibodies which include enzyme-linked immunosorbent assay (ELISA), lateral flow immunoassay (LFIAs) and chemiluminescent immunoassay (CLIAs). (5) Serological tests are helpful to identify asymptomatic and previously undiagnosed infections and thus are important in epidemiological surveys. Of particular importance are the neutralizing antibodies, which are capable of neutralizing the virus and thus provide protection against further infection. Plaque reduction neutralization test (PRNT) is the gold standard to detect the neutralizing capacity of the serum antibodies. The receptor-binding protein present in spike protein (S) of the virus interacts with human acetylcholine esterase-2 (ACE-2) receptor and thus helps the virus entry into the host cells. (6–8) Blocking the interaction between the S protein receptor-binding domain (RBD) and ACE-2 prevents the entry of the virus and thus is the most potent neutralizing epitope offering protection against SARS-CoV-2. (9,10)

The present study was conducted with the objective to measure the antibody level against SARS-CoV-2 among COVID-19 positive cases and to evaluate whether the presence of anti-SARS-CoV-2 antibodies indicates virus neutralizing capacity.

## Methodology

Present study was conducted among 100 participants who were enrolled from 15th March to 31 December 2021 from two sites, one rural site at Ballabgarh, Haryana, and another urban site at Dakshinpuri, New Delhi. Participation was voluntary. All the participants were recruited within 3 months of positive rapid antigen test report (RAT)/real time polymerase chain reaction (RT-PCR) report for COVID-19. Participants were enrolled into the study irrespective of their age or current COVID-19 disease status. Participants who refused to give written informed consent, or had contraindication for veni-puncture, were excluded from the study. From the consenting participants we collected information on basic demographic details, exposure history to COVID-19 cases, symptoms suggestive of COVID-19 in the preceding three month and clinical history.

Blood collection: Trained phlebotomists collected 5 ml of venous blood in plain vials from each participant within three months of testing positive for COVID-19. Serum was separated after centrifugation at the identified local health facility and transported to the respective laboratories for testing.

Detection of SARS-CoV-2-specific IgG antibodies was performed using an ELISA-based test (WANTAI) as per the specified optical density (OD) cut-off value. Neutralizing antibodies against SARS-CoV-2 was tested using plaque reduction neutralization test (PRNT) to check antibody titres. Anti-Receptor binding domain (RBD) antibody (IgG) was measured using quantitative RBD ELISA.

### Plaque Reduction Neutralization Test (PRNT)

PRNT for SARS-CoV-2 on Vero E6 cells was done to measure the neutralizing antibodies (SOP No.: THSTI/BL/TEC/039 Version: 1.0). PRNT_50_ was reported in titres. PRNT_50_ titre >20 was reported as positive, and PRNT_50_ titre of 20 or less was reported as negative. PRNT for SARS-CoV-2 had measurement uncertainty of ±19·14 at 936 PRNT_50_ titre of serum. (THSTI/BL/TEC/039: SOP for PRNT of SARS-CoV-2)

### Methodology of PRNT

The basic design of the PRNT assay allows virus-antibody interaction to occur in a microtiter plate, and then virus-antibody mixture was added on virus-susceptible cells. The antibody was subjected to serial dilutions prior to mixing with standardized amount of virus (i.e., 40 ± 20 PFU/well). The concentration of virus was kept constant such that, when added to susceptible cells and overlaid with semi-solid medium, individual plaques can be discerned when stained using crystal violet. Plaques were counted and compared with the virus controls to determine the percentage reduction in total virus infection.

The sensitivity of PRNT assay was defined as the ability of the assay to detect very low concentrations of a given substance in biological specimen. Sensitivity was defined as percentage of positive specimens out of already identified positives. Test was performed by one analyst on already ELISA tested positive samples (n=30). These samples were two-fold serially diluted starting 1:20 and the values were then compared. The observed results were compared to expected results. The sensitivity of the method was observed to be 100%. Specificity was defined as the ability of the assay to assess unequivocally the presence of components which may be expected to be present. It is also related to the concept of cross reactivity. Test was performed by one analyst on already ELISA tested negative samples (n=30). These samples were two-fold serially diluted starting 1:20 and the values were then compared. The observed results were compared to expected results. The specificity of the method was found to be 100%.

### QRBD

Quantitative enzyme-linked immunosorbent assay (ELISA) was used to estimate serum IgG antibodies binding to the receptor-binding domain of SARS-CoV-2 Spike protein (ELISA SOP No.: THSTI/BL/TEC/035, Version No. 1.1). The test reported the Anti-RBD IgG antibodies in ELU/ml. QRBD ≥ 12·0 ELU/ml was reported as positive, and between 8.0 and <12·0 ELU/ml was reported as equivocal. QRBD < 8·0 ELU/ml was reported as negative.

### Methodology for QRBD

The SARS-CoV-2 RBD IgG ELISA was done using a two-step incubation immuno-assay. Recombinant spike protein RBD antigen of SARS-CoV-2 was coated onto a 96-well polystyrene plate. The coated antigen can specifically recognize anti-RBD antibodies in human serum or plasma samples. After incubation, anti-RBD antibodies were captured by immobilized RBD protein while the unbound components were washed away. A detection solution containing HRP-conjugated anti-human IgG was added and plates were further incubated during which the HRP-conjugated anti-human IgG binds to the human IgG bound to RBD protein on the plate. After removal of nonspecific binding, a HRP substrate solution containing 3,3_′_,5,5_′_-Tetramethylbenzidine (TMB) was added, resulting in the formation of a blue color. Color reaction was stopped by 1Molar (M) sulphuric acid (H_2_SO_4_) which transforms the color of the solution from blue to yellow. The intensity of the color was quantified by measuring absorbance in a microplate reader at 450 nm. The color intensity represented directly the amount of anti-RBD antibodies captured inside the wells.

### WANTAI SARS-CoV-2 Antibody ELISA

It was an enzyme-linked immunosorbent assay (ELISA) for the qualitative detection of total antibodies to SARS-CoV-2 virus in human serum or plasma specimens (anti-SARS-CoV2 IgA, IgG and IgM antibodies). The kit is intended for screening of patients suspected of infection with the SARS-CoV-2 virus, and as an aid in the diagnosis of the coronavirus disease 2019 (COVID-19). Specimens with OD ≥ 0·19 were considered as positive, and <0·19 as negative.

### Methodology of WANTAI SARS-CoV-2 Ab ELISA test

WANTAI SARS-CoV-2 Ab ELISA was done using a two-step incubation antigen “sandwich” enzyme immunoassay kit, which used polystyrene microwell strips pre-coated with recombinant SARS-CoV-2 antigen. The patient’s serum or plasma specimen was added, and during the first incubation, the specific SARS-CoV-2 antibodies was captured inside the wells if present. The microwells were then washed to remove unbound serum proteins. Second recombinant SARS-CoV-2 antigen conjugated to the enzyme Horseradish Peroxidase (HRP-Conjugate) was added, and during the second incubation, the conjugated antigen bonded to the captured antibody inside the wells. The microwells were then washed to remove unbound conjugate, and Chromogen solutions were added into the wells. In wells containing the antigen-antibody-antigen (HRP) “sandwich” immune-complex, the colorless Chromogens are hydrolyzed by the bound HRP conjugate to a blue-colored product. The blue color turns yellow after the reaction was stopped with sulfuric acid. The amount of color intensity was measured and represented the amount of antibody captured inside the wells, and to the specimen respectively. Wells containing specimens negative for SARS-CoV-2 antibodies remained colorless. Specimens with an absorbance to Cut-off ratio of ≥ 1·0 were considered as positive.

### Statistical analysis

Categorical variables are reported as frequency and percentage. Normality of continuous variables was tested using Shapiro-wilk test. Continuous variables are reported as median with interquartile range. Wilcoxon ranksum test was applied to test the statistical significance of continuous variables. For testing correlation of categorical variables, Cramer V was calculated.

### Ethics

Ethical permission was taken from the institute ethics committee of All India Institute of Medical Sciences, New Delhi. (Ref. No.IEC-959/04.09.2020)

## Results

Mean (S.D.) age of the participant was 37·0 (13·5) years, the age ranged from 14 years to 72 years. Majority of the participants were male (64%).

Majority of the participants (63%) had history of fever, followed by cough (42%), sore throat (35%), and loss of taste sensation (24%). Seventy-four participants had at least one symptom, and the remaining 26 were asymptomatic.

Thirty-one participants had PRNT_50_ titre of less than 20, considered as negative for neutralizing antibodies.

Sixty-nine participants had neutralizing antibodies (PRNT_50_ titre ≥ 20). Among participants who had neutralizing antibodies (PRNT_50_ titre ≥ 20), 49 (71%) participants had PRNT_50_ titre ≥ 80, 37 (54%) had PRNT_50_ titre ≥ 160, and 26 (38%) had PRNT_50_ titre ≥ 320 (Figure 1) (Categories are not mutually exclusive).

**Figure 1:**
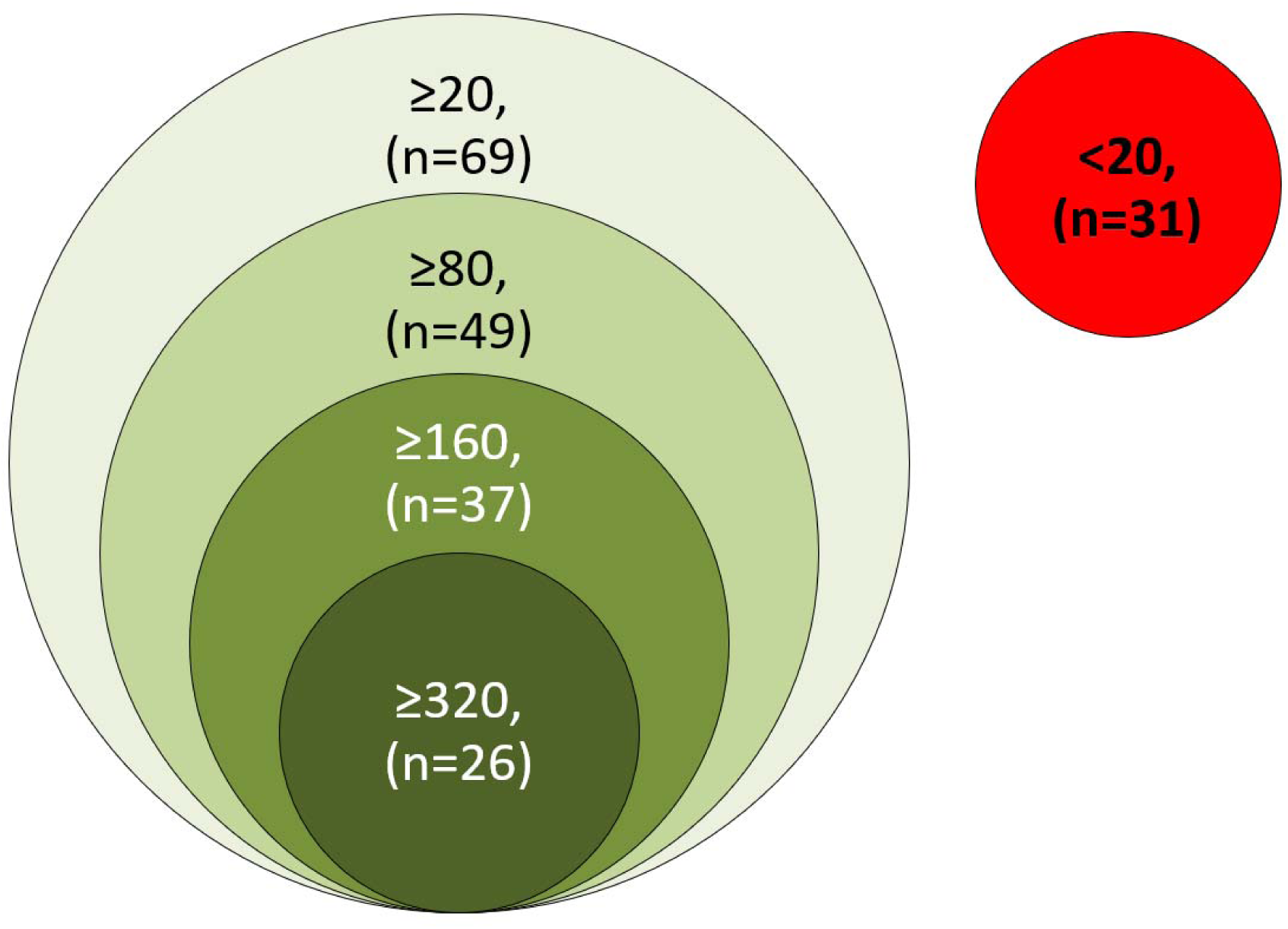
Venn diagram showing the PRNT_50_ titre of the study participants (<20 titre is considered negative, ≥20 is considered positive for presence of neutralizing antibodies).

Almost all participants (97·0%) were positive for Anti-RBD antibody (Serum IgG against receptor binding domain of COVID-19, done through QRBD) (≥ 12·0 ELU/ml), and three (3·0%) participant had equivocal result in QRBD (>7·99 to <12·0 ELU/ml). None of the participant was QRBD negative.

Among the 97 participants who were positive for anti-RBD, 69 (71·1%) had neutralizing antibody (PRNT_50_ titre ≥ 20). All the three participants with equivocal result for the QRBD, had PRNT_50_ titre of less than 20.

Almost all participants (99%) were positive for total anti-SARS-CoV2 antibodies (IgA, IgG, and IgM) (≥ 0·19 optical density (OD) in WANTAI assay.

The Cramer V (correlation coefficient) for presence or absence of receptor binding domain IgG (in QRBD) with presence/absence of neutralization antibody (PRNT_50_) was 0·2624 (p-value=0·028). It was even lower (Cramer V=0·1724, p-value=0·243) for presence or absence of receptor binding domain IgG with presence/absence of high titre of neutralization antibody (PRNT_50_ titre ≥ 80).

Mean (S.D.) PRNT_50_ (titre), and QRBD (ELU/ml) for the 100 participants were 543.1 (1341.5), and 465·0 (551·4) respectively. Median (IQR) PRNT_50_ titre was 71 (19, 415·5). Median (IQR) QRBD (ELU/ml) level was 202 (60, 627·6). Distribution of PRNT_50_ titre, and anti-RBD antibody levels showed non-normal distribution (Shapiro-wilk test, p<0·001, for both the variables). (Table 1)

**Table 1:**
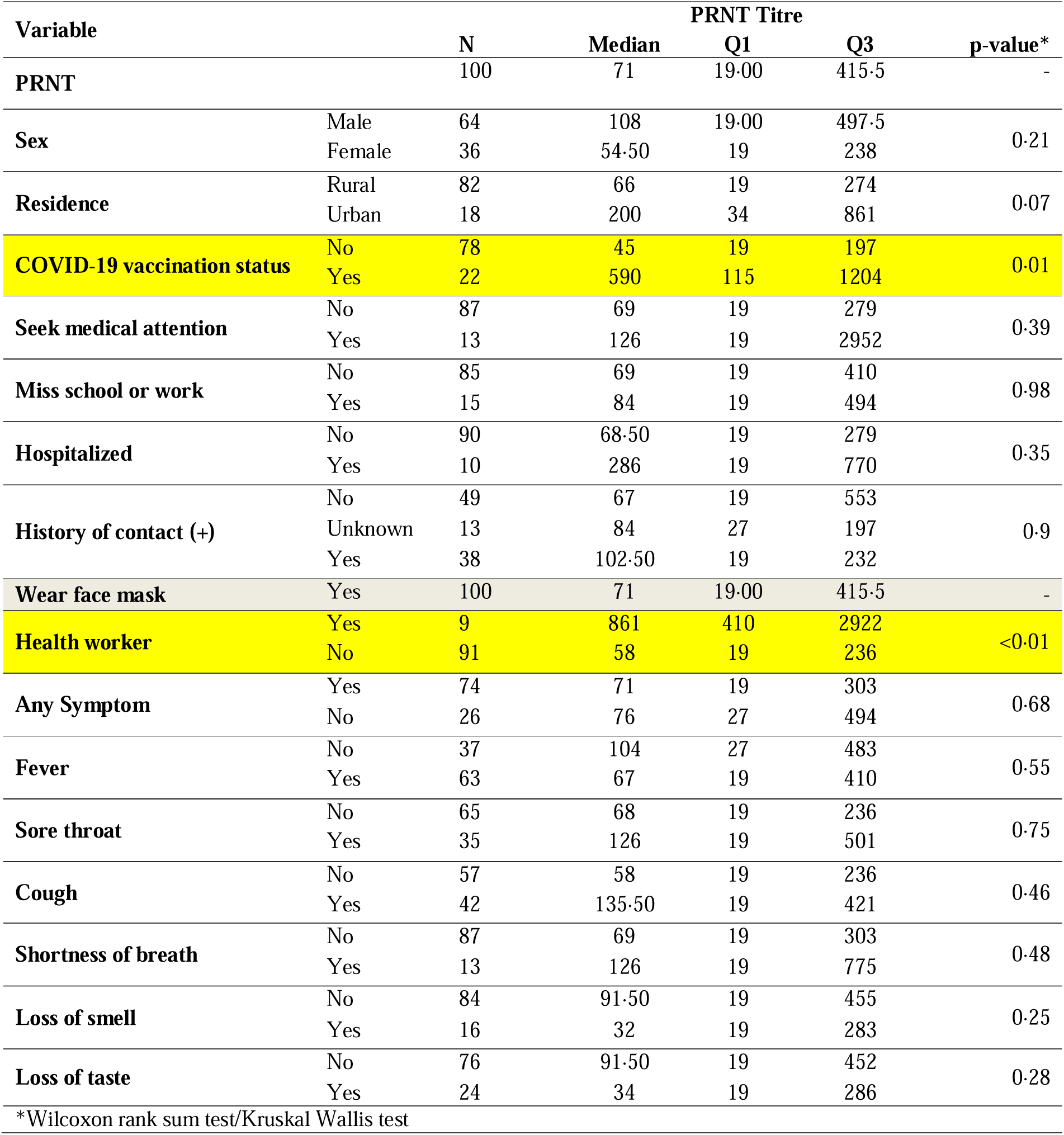
Distribution of PRNT titre by sociodemographic and clinical variables.

Figure 2 and 3 show the distribution of the PRNT_50_ (titre), and QRBD (ELU/ml) for the 100 participants. Figure 4 shows the scatter plot of Anti-RBD antibodies (ELU/ml) (QRBD) among the participants with Neutralising antibody titre (PRNT) among them.

**Figure 2:**
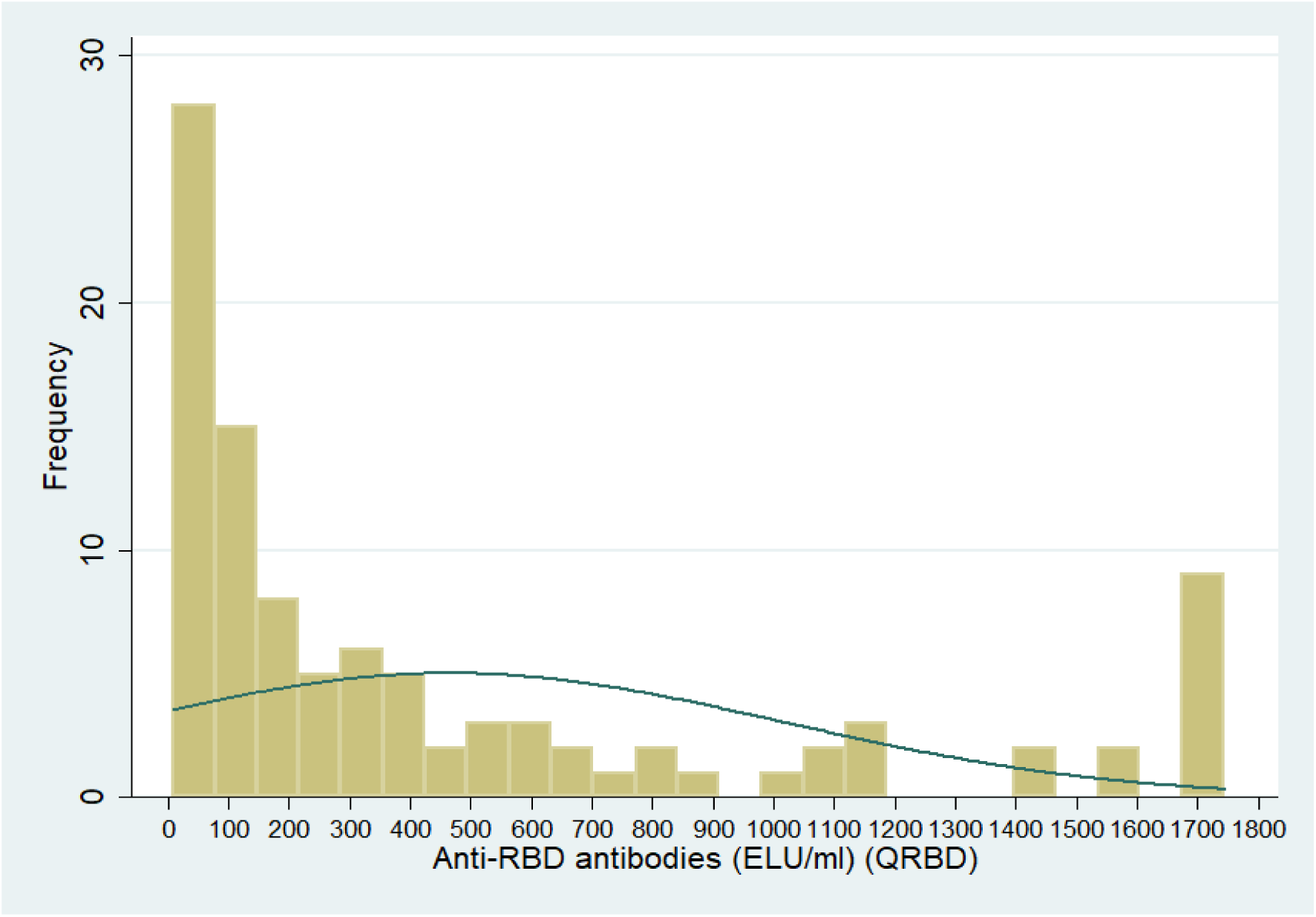
Distribution of Anti-RBD antibodies (ELU/ml) among study participants.

**Figure 3:**
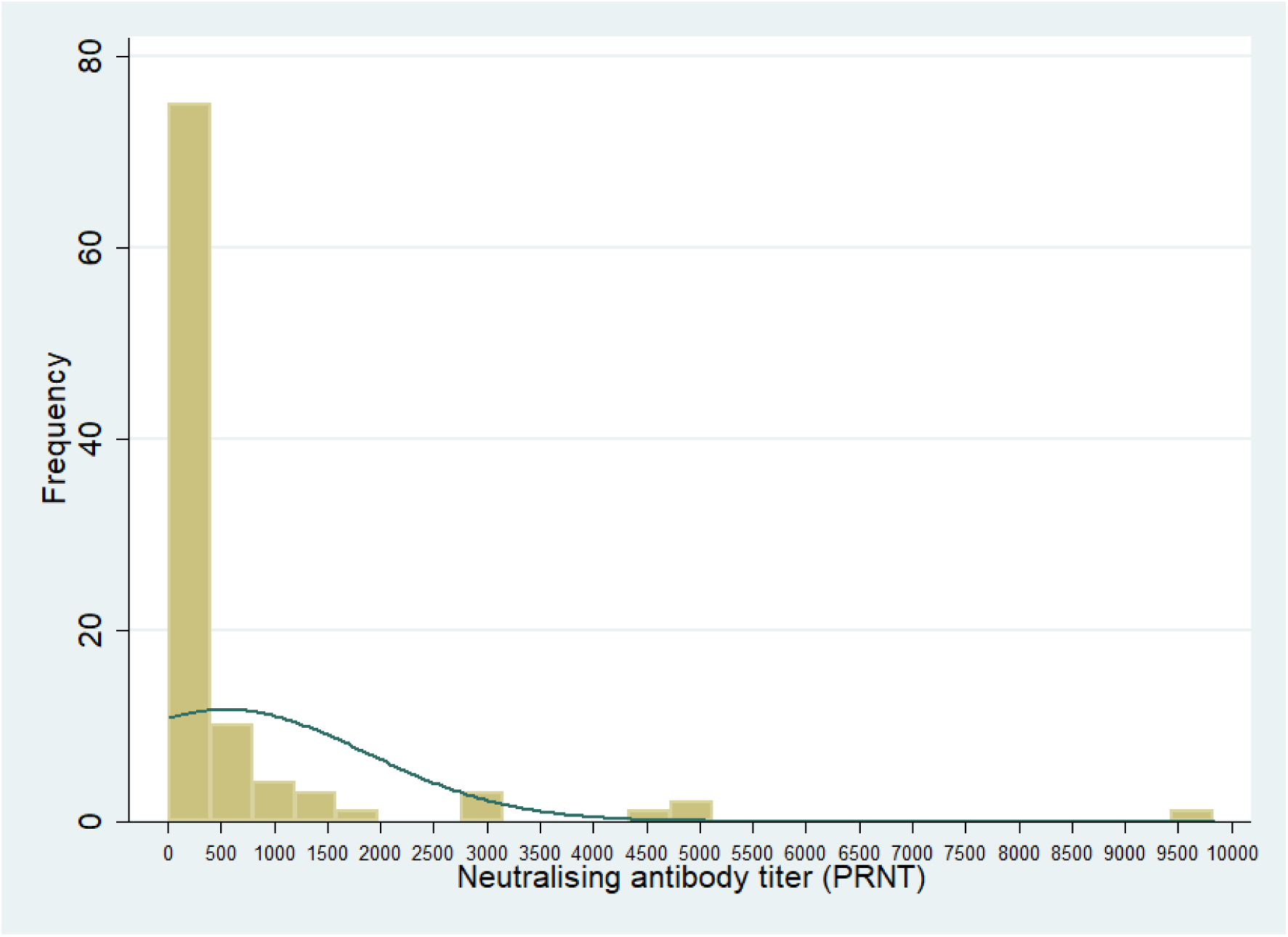
Distribution of Neutralising antibody titre among the participants.

**Figure 4:**
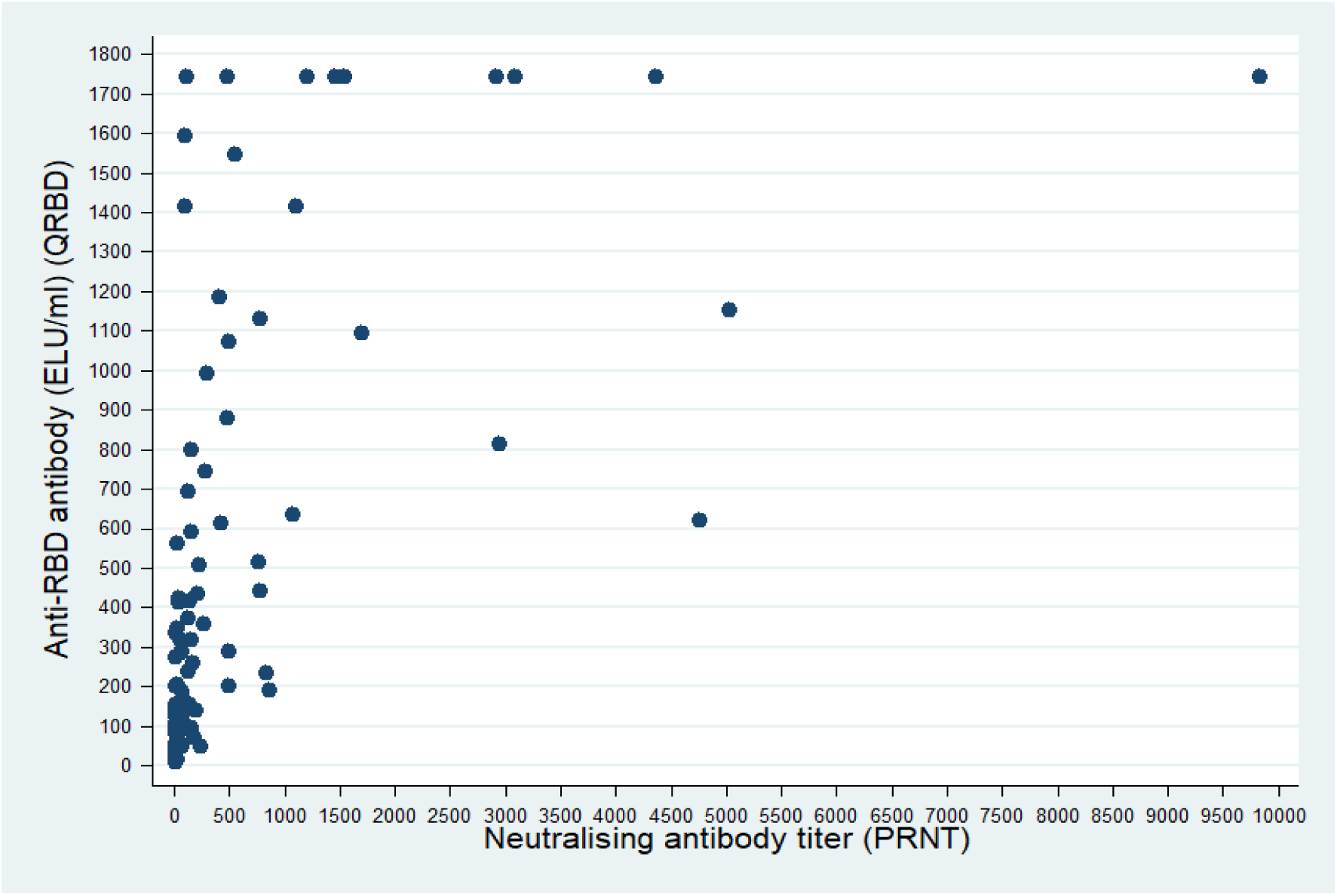
Scatter plot of Anti-RBD antibodies (ELU/ml) among participants with Neutralising antibody titre among them.

Median PRNT_50_ titre was higher among males compared to females, among urban residents compared to rural, among participants who got at least one dose of COVID-19 vaccine compared to those who didn’t, among those who sought medical attention compared to those who didn’t, among participants who were hospitalised compared to who didn’t, and among health care workers compared to other occupations. Median PRNT_50_ titre was higher among participants who had symptoms of sore throat, cough, and shortness of breath, compared to those who didn’t. However, participants with loss of smell, and loss of taste had lower median PRNT_50_ titre. Median (IQR) PRNT_50_ titre was significantly higher among those who received at least one dose of COVID-19 vaccine compared to no vaccination [590 (115, 1204) vs. 45 (19, 197), Wilcoxon rank sum test, p-value =0.01]. Similarly, Median (IQR) PRNT50 titre was significantly higher among participants who were working as health care workers compared to those who weren’t [861 (410, 2922) vs. to 58 (19, 236), Wilcoxon rank sum test, p-value<0·01]. For other variables, the difference was not statistically significant. Table 1, shows the median (IQR) PRNT_50_ titre with respect to different sociodemographic/clinical variables.

Median (IQR) Anti-RBD antibody level was significantly higher among residents of urban area compared to rural area [437·6 (141·9, 1183·4) vs. 192·1 (55·4, 589·8), Wilcoxon rank sum test, p-value =0·04]. Those who had taken at least one dose of COVID-19 vaccine had significantly higher Median (IQR) anti-RBD antibody level compared to unvaccinated [718.1 (441.3, 1415.9) vs. 131 (52·3, 372·3), Wilcoxon rank sum test, p-value <0·01]. Participants who had history of contact with COVID positive cases had significantly higher median (IQR) QRBD titre of 348·2 (126·8, 1094·8) compared to those who had no history of contact with COVID positive cases (128·2 (48·7, 414·8)) (Wilcoxon rank sum test, p-value <0·01). Participants who were working as a health worker also had significantly higher median (IQR) QRBD titre compared to those who were not health worker [798·4 (441·3, 1415·9) vs. 184·7 (55·6, 560·7), Wilcoxon rank sum test, p-value <0·01].

Similar distribution as for PRNT titre were seen for Anti-RBD antibody levels with respect to sociodemographic/clinical variable, except for two variables: (a) History of contact: median PRNT_50_ titre was higher amongst those exposed, while median anti-RBD antibody levels were higher among those not exposed; (b) Presence/absence of fever as symptom: median PRNT_50_ titre was higher among those who had no fever, while median Anti-RBD antibody levels were higher among those reporting fever. Among the participants with anti-RBD antibodies: the anti-RBD antibody levels were significantly higher for participants that had neutralizing antibodies compared to those who didn’t have neutralizing antibodies (Wilcoxon rank sum test: p-value <0·001). Overall, also the anti-RBD antibody levels were significantly higher among participants who had neutralizing antibodies compared to participants who do not have neutralizing antibodies (Wilcoxon rank sum test: p-value <0·001). (Table 2) (Figure 5)

**Table 2:** Distribution of anti-RBD antibody levels by Neutralizing antibody titres status.

| Anti-RBD antibody status | PRNT<sub>50</sub> titre status |  |  |  |  |
| --- | --- | --- | --- | --- | --- |
|  | Negative (PRNT<sub>50</sub><20) |  | Positive (PRNT<sub>50</sub>≥20) |  | Wilcoxon rank sum test (p-value) |
|  | Frequency (%) | Anti-RBD antibody levels (ELU/ml) Median (IQR) | Frequency (%) | Anti-RBD antibody level s (ELU/ml) Median (IQR) |  |
| Positive | 28 (28%) | 53.8 (34.0, 116.75) | 69 (69%) | 414.8 (169.2, 1073.7) | <0.001 |
| Equivocal | 3 (3%) | 11.7 (9.1, 11.9) | 0 (0) | - | - |
| Total | 31 (31%) | 48.7 (25.0, 107.4) | 69 (69%) | 414.8 (169.2, 1073.7) | <0.001 |

**Figure 5:**
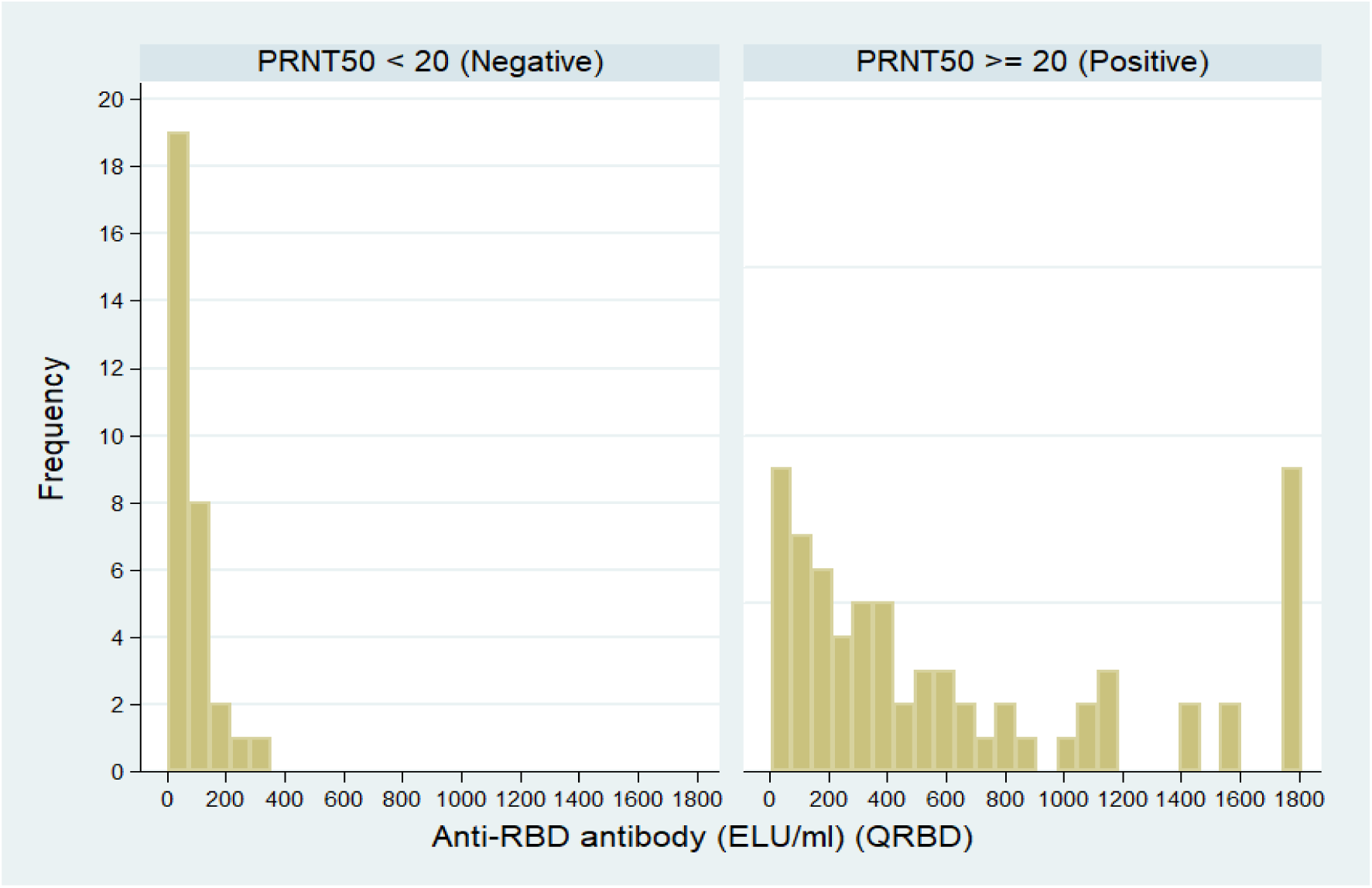
Histogram showing Anti-RBD antibodies (ELU/ml) among the participants with respect to PRNT_50_ <20 and PRNT_50 ≥_ 20.

## Discussion

Present study was conducted among one hundred laboratory confirmed COVID-19 positive cases. All participants were tested by PRNT, WANTAI and QRBD for COVID-19 neutralizing antibodies, total antibodies (IgA, IgG, and IgM) against COVID-19, and Anti-RBD IgG antibodies for COVID-19 respectively.

Though almost all participant had anti-RBD IgG antibodies (97% positive, 3% equivocal result), and anti-COVID-19 antibodies (IgA, IgG, and IgM) (99%), only sixty-nine (69%) participants had neutralizing antibodies. Therefore, just the presence of Anti-SARS-CoV-2 antibodies doesn’t mean that the person has neutralizing antibody titre and thereby protected against the virus.

In the study by Deshpande et al (11) among 343 participants, 71·9% developed neutralizing antibodies to SARS-CoV-2. Among the 28·1% (n=25) participants failed to develop neutralizing antibodies; eleven participants were positive by anti-SARS-CoV-2 IgG ELISA. The participants in their study differed from our study. We had included only laboratory confirmed COVID-19 cases, however their study sample consisted mixed sample (89 positive, 58 negatives for SARS-CoV-2 and 17 cross-reactive and 179 serums from healthy participants). Also, they reported PRNT_90_ instead of PRNT_50_ as was the case in this study. The difference noted, therefore, could be due to difference in methods.

Lau et al (12) reported that 99·1% of the participants had neutralizing antibodies at 90 days after symptoms/detection of infection, in the serum sample from 195 RTPCR positive cases of COVID-19. The difference could be because of difference in the disease spectrum of the recruited patients. The study by Lau et al had only 31 asymptomatic cases (15%), while in present study 26% individuals were asymptomatic.

Although previous studies have shown good correlation between the Anti-RBD antibodies and virus neutralization (13,14), present study shows a different picture as almost all the participants had Anti-RBD antibodies, but only 69% had neutralizing titre. Anti-RBD antibody levels were significantly higher among participants that had neutralizing antibodies compared to participants who didn’t. It shows that there is need to increase the cut-off point of the anti-RBD antibody levels which then can act as a proxy indicator for presence of neutralizing antibodies.

Also, in present study only 41% of the participants had high titre of neutralizing antibodies (PRNT_50_ titre≥ 80).

Neutralizing antibody titre, and Anti-RBD antibodies were significantly higher among vaccinated even if with one dose of COVID-19 vaccine. It therefore, appears that vaccination provides protection against COVID-19. Our finding is in agreement with the other previous studies. (14,15)

Also neutralizing titre and anti-RBD antibody titre were significantly higher among the health care workers, which might be due to repeated exposure of SARS-CoV2 among them. Participants who had history of contact with COVID-19 positive participants had significantly higher anti-RBD antibodies. Urban residents had significantly higher anti-RBD antibodies compared to rural, which could be due to higher population density in urban areas and thereby higher probability of repeated exposure to SARS-CoV-2.

### Strengths

Only laboratory confirmed cases of COVID-19 were recruited in the study. We had measured neutralizing antibodies through PRNT assay which is considered gold standard. Also, all the sera samples were tested for Anti-RBD antibodies and total antibodies. Standard kits and protocol were followed for all the assays. All the sera samples were taken within 3 months of positive RT-PCR/RAT tests.

### Limitations

Symptoms and history of contact were self-reported hence making it vulnerable to recall error.

## Conclusion

Almost all the participants had Anti-SARS-CoV2 antibodies (IgG and IgM) and Anti-RBD IgG antibodies. However only 69% had neutralizing antibodies against SARS-CoV2. Proportion of participants with higher titres of neutralizing antibodies was even lower, almost 50%. The present study highlights that presence of antibodies against SARS-CoV2, or presence of anti-RBD antibody doesn’t necessarily imply presence of neutralizing antibodies.

## Data Availability

Deidentified patient data will be made available with investigator support, with a signed data access agreement.

## Acknowledgement

We acknowledge World Health Organisation for providing support for this study.

